# Patient and public experience and views on digital systems for sharing records for health and care preferences at the end of life

**DOI:** 10.1101/2024.06.19.24309169

**Authors:** J. Birtwistle, AM Russell, S. Relton, H. Easdown, U. Grieve, M. Allsop

## Abstract

**Objective:** To explore patient and public experiences of and priorities for the use of shared patient health records for advance care planning.

**Methods:** A convergent-parallel mixed method design was used. An online national survey of patients and the public gathered data on experiences and views of sharing health and advance care planning information to support care at the end of life. Descriptive statistics were used to analyse rating scale responses (5 or 10-point scale) and thematic analysis applied to free-text responses.

**Results:** Responses (N=1728) included participants in 103 UK counties, including people with a terminal condition (n=33), long-term condition (n=442), who provide or have provided care to a person with a long-term or terminal illness (n=229), and who identified as healthy and interested in planning for the future (n=1024). Confidence that recorded care preferences would be accessed when needed was low for carers (median= 2, IQR 1-4) and moderate for patients (median=3, IQR 1-4). Four themes derived from free-text responses included: i) Experience of sharing health information; ii) Preparation, communication and understanding; iii) Concerns, unknowns and assurance seeking, and; iv) Preserving Dignity and Respect: Understanding individual contexts.

**Conclusions:** Whilst recognising the potential of sharing health records, respondents and in particular carers, doubted that patient information would be accessed by relevant health professionals when needed. Future research is required to explore whether patient and carer access to the record influences their confidence in the accuracy of the content and the likelihood of care being delivered in line with their wishes.

**What is already known on this topic:** Digital systems can support documentation and sharing of health information, wishes and preferences for the end of life.

**What this study adds:** Patients and carers perceive the documentation of advance care plans as a burdensome and complex process, that can lead to confusion about the purpose of documentation.

Respondents expressed doubts about the accuracy of documented information, uncertainty about whether health professionals could access their records when needed and concerns that documented wishes and preferences would be ignored.

**How this study might affect research, practice or policy:** Patient and public views must be considered in the design and implementation of digital systems. In particular, efforts should be made to build confidence and clarify the expectations of patients and members of the public around the documentation of their wishes and preferences for care alongside the subsequent sharing and use of this information.

## Background

Advance care planning involves supporting people in understanding and documenting their values and preferences regarding future medical care. Creating and documenting an advance care plan for patients with long-term or life-limiting conditions can help align care at the end of life with the patient’s wishes(1). The success of documenting this plan often depends on the extent to which patients can articulate their values and identify which treatments would align with these in future scenarios, and that clinicians can elicit and document these values and preferences(2). A further step to success supposes that clinicians will access, read, and honour the patient’s expressed wishes and that health and care services commit resources to support preferred patient care(2).

Depending on the country context and resources available, patients receiving palliative care may require input from many health professionals in various settings including community healthcare, hospital services, emergency services, and residential care(3). Documentation of preferences for care may need to be recorded, viewed, updated, and acted upon by health professionals involved in delivering palliative and end-of-life care and working in these different health and care settings(4). Increasingly, digital systems are being developed across health and social care providers in the statutory, voluntary and private sectors, aiming to provide more efficient information-sharing, reduce duplication of work, and enable health professionals to spend less time finding patient information(4-6). Digital systems for supporting documentation of care preferences typically take the form of a template in an electronic patient health record or a standalone online system that can be linked to a health record(6). Where an advance care plan is recorded for patients with a terminal or progressive illness, these systems aim to make the information available across all settings involved in a person’s care (e.g. specialist hospital services, care homes and hospices)(7-9). The information held may reflect conversations held at different times with different health professionals. There are, however, known issues with the implementation of systems in some settings such as the UK, where interoperability of multiple electronic health record systems prevents information sharing across all settings alongside electronic and paper-based approaches running in parallel(10-12).

Alongside digital platforms for information sharing by health professionals, online resources are increasingly being used to support advance care planning decisions and documentation, without direct input from a health professional(13). Patients often complete these independently or with an advocacy service(13). These resources typically require the download of the patient-completed form that must be taken to a health professional to be replicated or a copy made and uploaded to a digital health record(14, 15). These resources have been shown to increase engagement with the advance care planning process but are standalone documents and not linked with digital systems used by health professionals(16).

Whilst a growing number of studies point to the patient experience of digital sharing of patient health information, specifically to patient online access (e.g. (17-21)), there is a lack of research into how using systems for documenting, sharing and accessing advance care planning information contributes to patients’ experience(22). A crucial missing element from the evidence base is the voice of patients and caregivers who have been largely excluded from the design and development of digital systems for sharing advance care plans.

## Methods

### Aim

This research aimed to explore patient and public experiences and views on digital systems for sharing health and care preferences at the end of life. We sought to understand participants’ perceptions of information sharing and its use for patient care at the end of life, and how the use of digital systems might influence levels of confidence around the accuracy and use of documented information.

### Design

A data analysis of an online survey using a convergent-parallel mixed method design.

### Setting

As part of a comprehensive public consultation, Compassion in Dying and the Professional Record Standards Body (PRSB) undertook a national survey to seek views from patients, carers and members of the public in the UK. The survey focused specifically on advanced care planning and digital care planning which includes Electronic Palliative Care Coordination Systems, commonly abbreviated to ‘EPaCCS’. EPaCCS are digital systems that have been developed in England that aim to facilitate the documentation of advance care planning information for health conditions requiring multi-professional and multi-setting involvement, communication, and coordination around expressed patients’ wishes (7). This is the first comprehensive analysis and reporting of this survey data.

### Participants

A convenience sample was used. Individuals could be included if they self-identified as having a terminal condition, a long-term condition, being a current or bereaved carer of a person with a terminal or long-term condition or a healthy person interested in planning for their future care.

### Data collection

Data were collected in February 2021 using Survey Monkey (http://www.surveymonkey.com). The survey link was distributed via email by Compassion in Dying and the PRSB. Emails were sent to around 18,500 people held on a confidential register containing people who had been supported by Compassion in Dying since 2016 and agreed to hear about new content or events. No reminder emails were sent. The survey was advertised in the PRSB newsletter (around 2850 subscribers) and on social media (Twitter (X): around 2750 followers). NHS England asked members of their digital workstream group to share the survey link with organisations they represent. This comprised hospices, clinicians, and end-of-life charities. The survey was open for 13 days.

### Survey

The survey questionnaire was designed and tested for face validity and readability in consultation with patients and carers who work together to provide oversight and guidance to NHS England’s strategic programmes. Patients and carers provided feedback on language, clarity and how questions about end-of-life wishes could affect the people completing the survey. Questions were adapted according to recommendations. Four questions sought information on experiences and views about documentation and sharing health records within and between different services. Specifically, they asked about recording, sharing and accessing wishes and preferences at the end of life. Participants were asked to specifically think about if they or a loved one became more ill and needed to be cared for somewhere else. Questions, including four free-text comment boxes, are shown in Supplemental File 1. Demographic information included the role of the respondent and county of residence. Anonymised data were transferred from Compassion in Dying to the researchers using encrypted software. Participants were free to choose whether to complete the survey. We report the study in accordance with the Checklist for Reporting Results of Internet E-Surveys (CHERRIES) guideline for on-line survey distribution and reporting(23).

### Ethics

Ethical approval was granted by the University of Leeds, School of Medicine (MREC 21-032) to undertake a comprehensive analysis of the anonymised data.

## Analysis

### Quantitative analysis

Descriptive statistics were used to analyse statement ratings for confidence in health professionals’ access to health information and features of records that would increase confidence in end-of-life care provision (medians and interquartile range); options for permission to share were analysed using counts and proportions. A Mann-Whitney U-test was used to explore whether respondents’ level of confidence that health professional would access their records had any bearing on their likelihood of entering a comment about their views or experience.

### Qualitative analysis of free-text comments

A dual analysis was adopted for data generated through the free-text items in the survey.

#### Comment boxes 2 and 3

Free-text comments relating to questions in boxes 2 (*We would like to know how you feel about information about your <u>health</u> being shared with your healthcare team*) and 3 (*We would like to know how you feel about your <u>end-of-life wishes and preferences</u> being shared with your healthcare team*) were analysed using conventional content analysis(24). These comment boxes were presented with their respective question as an alternative to a fixed response. Comments were checked for unique content and summarised descriptively with each question.

#### Comment boxes 1 and 4

A thematic approach was used for responses in comments boxes 1 (*If there’s something else in relation to your end-of-life care record that is important to you, please tell us here*) and 4 (*If you’ve had a specific experience, or have thoughts about your wishes being known at the end of life, could you tell us a bit more about it?*). This method was considered appropriate to capture and describe the diversity of perspectives and experiences reported (25, 26). Using a structured approach(27), data were divided into responses for each respondent with comments in both boxes (1 and 4) considered as a whole response. One researcher (JB) familiarised herself with the data for each respondent by reading all the responses. When creating themes, JB reviewed coding and themes with AR and MA and revised them based on group agreement.

## Results

There were a total of 1728 responses from eligible participants from most UK counties (103/109; 94.5%) including people with a terminal condition (n=33), a long-term condition (n=442), who provide or have provided care to a person with a long-term or terminal illness (n=229), and people who identified as healthy and interested in planning for the future or offered other reasons for completing the survey (typically a mild condition or a carer for someone with one) (n=1024). Four of these people with a terminal illness and 70 with a long-term condition also provide or have provided care to a loved one with a long-term or terminal illness. Missing data were low at < 2% for most questions. All medians are calculated from responses provided (i.e. excluding missing data).

### Confidence that the healthcare team will have access to the information they need to support or treat the patient at the end of life

Respondents’ ratings of confidence that the healthcare team supporting or treating them will have access to information about them when caring for them (or the person they care for) at the end of life are shown in Table 1. Median ratings indicated a moderate level of confidence that health professionals will access their end-of-life information, this was similar across respondent role groups and the type of information accessed.

**Table 1:** Median ratings of level of confidence in health professional access to respondents’ information at the end of life.

| **The healthcare team supporting or treating me or a loved one will have access to:**<br>*Please rate how confident you feel about the following statements*<br>*(1 to 5 scale, where 1 = not confident at all and 5 = very confident):* |  |  |  |  |  |  |  |  |  |  |  |  |
| --- | --- | --- | --- | --- | --- | --- | --- | --- | --- | --- | --- | --- |
|  | **Terminal condition (N= 33)** |  |  | **Long term condition (N=442)** |  |  | **Carer (N =229)** |  |  | **Healthy/other (N=1024)** |  |  |
|  | *n* | *Mdn (IQR)* | *DK n (%)* | *n* | *Mdn (IQR)* | *DK n (%)* | *n* | *Mdn (IQR)* | *DK n (%)* | *n* | *Mdn (IQR)* | *DK n (%)* |
| Information about my health that they need when caring for me at the end of life | 30 | 3 (1-4) | 3 (9) | 386 | 3 (1-4) | 53 (12) | 214 | 3 (1-4) | 14 (6) | 871 | 3 (2-4) | 145 (14) |
| My end-of-life wishes and preferences | 26 | 3 (1-4) | 7 (21) | 380 | 3 (1-4) | 58 (13) | 208 | 2 (1-4) | 17 (7) | 879 | 3 (1-4) | 138 (13) |
| Information on who I want to be involved in decisions about my health and care at the end of life | 25 | 3 (1-4) | 7 (21) | 387 | 3 (1-4) | 51 (12) | 213 | 2 (1-4) | 15 (7) | 885 | 3 (1-4) | 134 (13) |

Medians calculated as those who provided a rating (i.e., discounting missing data and respondents who answered "Don't know")
Key: DK Don't know

### Priorities for increasing confidence that the healthcare team will have access to the information they need to support or treat the patient at the end of life

Respondents were asked to rate aspects of digital systems on their ability to increase confidence that they or a loved one would get the care that’s right for them or the person they care for at the end of life (Table 2). Ratings were similar across participant role groups and items. Median ratings indicated the most highly rated aspects were that health professionals have access to preferences, including the details of the people they want to be involved in care decisions and preferences, such as the place of care and treatments they do and don’t want. There was also a high level of importance placed on the ability for patients access to their digital records with the ability to view, edit, and share with family.

**Table 2:**
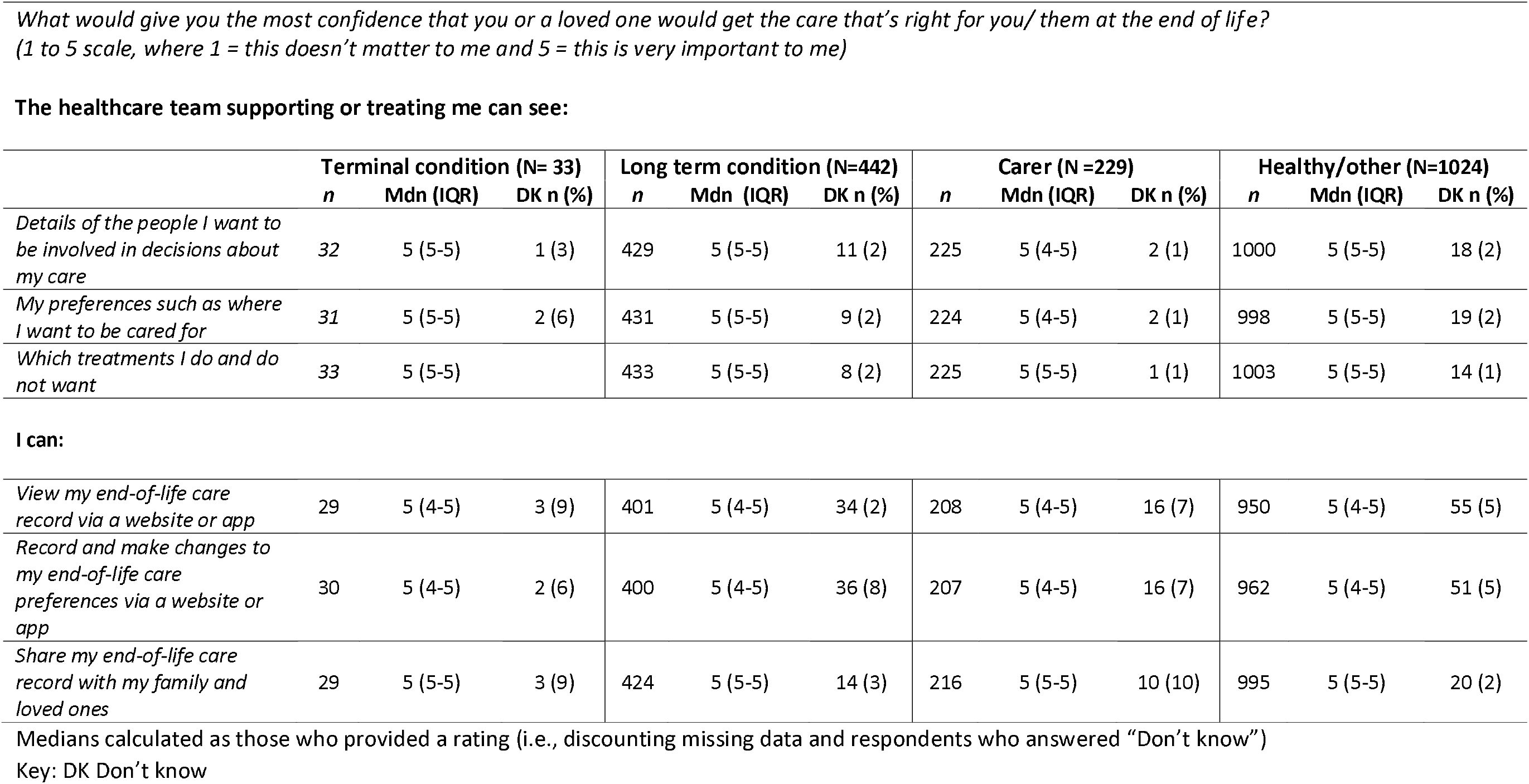
Median ratings of confidence in features to support health professionals’ provision of end-of-life care.

**The healthcare team supporting or treating me can see:**
|  | Terminal condition (N= 33) |  |  | Long term condition (N=442) |  |  | Carer (N =229) |  |  | Healthy/other (N=1024) |  |  |
| --- | --- | --- | --- | --- | --- | --- | --- | --- | --- | --- | --- | --- |
|  | *n* | Mdn (IQR) | DK n (%) | *n* | Mdn (IQR) | DK n (%) | *n* | Mdn (IQR) | DK n (%) | *n* | Mdn (IQR) | DK n (%) |
| *Details of the people I want to be involved in decisions about my care* | 32 | 5 (5-5) | 1 (3) | 429 | 5 (5-5) | 11 (2) | 225 | 5 (4-5) | 2 (1) | 1000 | 5 (5-5) | 18 (2) |
| *My preferences such as where I want to be cared for* | 31 | 5 (5-5) | 2 (6) | 431 | 5 (5-5) | 9 (2) | 224 | 5 (4-5) | 2 (1) | 998 | 5 (5-5) | 19 (2) |
| *Which treatments I do and do not want* | 33 | 5 (5-5) |  | 433 | 5 (5-5) | 8 (2) | 225 | 5 (5-5) | 1 (1) | 1003 | 5 (5-5) | 14 (1) |
| **I can:** |  |  |  |  |  |  |  |  |  |  |  |  |
| *View my end-of-life care record via a website or app* | 29 | 5 (4-5) | 3 (9) | 401 | 5 (4-5) | 34 (2) | 208 | 5 (4-5) | 16 (7) | 950 | 5 (4-5) | 55 (5) |
| *Record and make changes to my end-of-life care preferences via a website or app* | 30 | 5 (4-5) | 2 (6) | 400 | 5 (4-5) | 36 (8) | 207 | 5 (4-5) | 16 (7) | 962 | 5 (4-5) | 51 (5) |
| *Share my end-of-life care record with my family and loved ones* | 29 | 5 (5-5) | 3 (9) | 424 | 5 (5-5) | 14 (3) | 216 | 5 (5-5) | 10 (10) | 995 | 5 (5-5) | 20 (2) |

Medians calculated as those who provided a rating (i.e., discounting missing data and respondents who answered "Don't know")
Key: DK Don't know

### Preferences for sharing records with health professionals

The majority of respondents (68-75% across all role groups) said they would like their information about health and end-of-life wishes and preferences to be available to any healthcare professional supporting them whenever they need it, without having to give their permission for this information to be shared (Fig.1). A further 23-31% said they would prefer to be asked once for permission. Respondents who chose neither of the above options were invited to provide further information about their preference for access to their information. Content analysis of 82 comments indicated other preferences for sharing information. Preferences included that permission should be sought from the next of kin at the point of need, and should be situation- or service-specific only (e.g. permission-free access for emergency situations or health professionals already known to the person).

**Figure 1.**
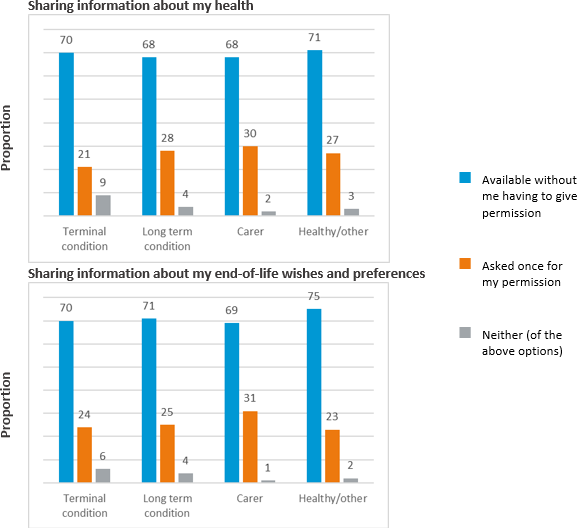
Preferences for sharing information with health professionals. Percentages calculated as those who responded (i.e.. discounting missing data)

### Qualitative Data

Of the 1726 survey participants, 1071 entered a free text response in at least one comment box (boxes 1 or 4). The number of participants who commented and the types of responses coded are shown in Table 3. Comments from the healthy/other group (556 respondents) were excluded from free-text analysis to specifically look at those with terminal illness and long-term conditions.

The Mann-Whitney U-test found that respondents who left at least one comment reported significantly lower levels of confidence that the healthcare team supporting or treating them could see their information than those who left no comment. This was true for all types of information: Information about my health that they need when caring for me at the end of life (U = 50548.0, p = 0.0171); My end-of-life wishes and preferences (U = 48481.5, p = 0.010) and Information on who I want to be involved in decisions about my health and care at the end of life (U = 52302.5, p = 0.0005).

Participants in all groups often spoke from the viewpoint of both carer and patient (e.g. patients also spoke about experiences as a carer; carers often considered their own end-of-life care as well as the person they care for). How people experienced, made sense of, and stated their planning needs, discussing, documenting, and sharing end-of-life wishes and preferences is captured in four themes: i. Experience of sharing health information; ii. Preparation, Communication and Understanding; iii. Concerns, unknowns and assurance seeking; iv. Preserving Dignity and Respect: Understanding individual contexts. The conceptual map (Fig. 2) considers the digital documentation and sharing of wishes and preferences in detail. It shows emerging themes that describe how the recording process and the digital system interact with patients’ needs and concerns and how they perceive it.

**Figure 2:**
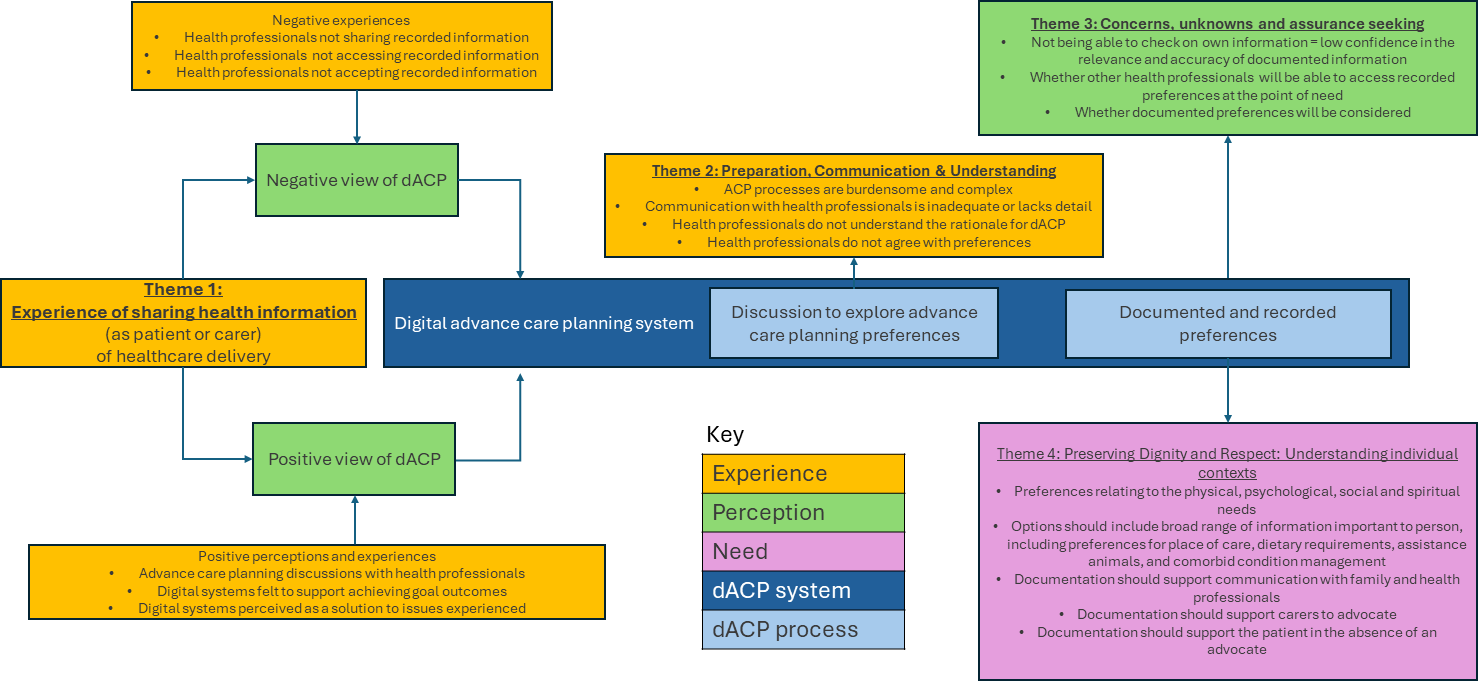
Conceptual map of digital documentation and sharing wishes and preferences.

### Qualitative Analysis

#### Theme 1: Experience of sharing health information

Prior experience of sharing medical or advance care planning information, whether it was digitally transmitted or via other means, shaped participants’ views about how digital records could support their needs at the end of life. Experiences reported included observations relating to the care of another person as well as participants’ own experiences.

Respondents who talked about the perceived benefits of digitally recorded advance care plans had either experienced a positive encounter related to discussion, documentation or sharing end-of-life wishes or had reported an issue. Access to a digital system was perceived as a solution to issues experienced with sharing patient wishes within organisations (Carer 815) and between them (Carer 321).

> *“Every time I go to hospital with my elderly mother, they lose her advanced directive or fail to pass it from one ward to another. A recognised electronic form would be fantastic and very reassuring.”* **Carer 815**

> *“I am aware that her DNAR requests are logged with her GP and should be also with the hospital but it seems this is not shared with the hospital. To have the DNAR available online with access allowed would be a benefit.”* **Carer 321**

They perceived that health professionals are working together to plan, document and share patient information and that they would access patients’ end-of-life wishes at the point of need.

> *“Easy to have a conversation with my GP about my wishes. She recorded my wishes on the computer system, and has contacted me every 6 months to ensure there is no amendment.”* **LTC 828**

Conversely, participants who held a negative perception of documentation or sharing of their information had often experienced instances where health professionals had not accessed their information or had been reluctant to accept or action it.

> *“I am in my 30s and have a DNACPR, RESPECT form and Advanced Directive in place due to serious health complications. I have often had Ambulance staff and hospital staff not believe my story.”* **Long Term Condition (LTC) 163**

#### Theme 2: Preparation, communication and understanding

Respondents were keen to document their advance care plan so it would be available to health professionals. However, they considered this to be a burdensome, complex process they would have to complete alone or with inexperienced health professionals.

> *“I would really like to crack on and get this sorted, H&W PoA, Advance Directives etc. but the bureaucracy is daunting.”* **LTC 1525**

> *“I have completed the Coordinate My Care process, which shares info from GP with the ambulance service. It is not simple yet rather limited, basically DNAR? and do you want to die at home? My GP had never done this before.”* **LTC 444**

Communication about what advance care planning involves was often inadequate and lacked detailed information about the process that the patient or family could understand. This led to confusion for carers about the purpose of this discussion and a lack of confidence in health professionals’ understanding of their family members’ wishes.

> *“GP offered a discussion about DNACPR and ceilings of treatment by sending a post-it note home with a family member after a routine appointment. Family members didn’t know what this was. Not well explained…..Not really confident in GP’s understanding of Dad’s wishes.”* **Carer 142**

They reported discrepancies between their own and health professionals’ understanding of their rationale for documenting their preferences and as well as the decisions they have made, and often felt their ideas and preferences would not be accepted by health professionals and therefore ignored.

> *“I have given my GP my ACP, but she seemed perplexed as to why I was doing this since I was not ill. I am not convinced they really get the importance of doing this early, basically because I think many medical workers are still stuck with the idea that their job is about curing us, not helping us to live well.”* **LTC 671**

#### Theme 3: Concerns, unknowns and assurance seeking

Respondents expressed concerns about health professional interaction with their information at each stage of digital documentation of preferences. They raised concerns about the level of accuracy of the information recorded about them, and they reported times when health professionals could not easily access their information. Additionally, they were unable to check their own information, resulting in little confidence that it would be accurate and relevant. The following quote describes the person’s perception of the use of paper verses digital records for their data and the unmet need for the patient to check the information recorded.

> *“Many professionals have been involved in my relative’s care. Most bring & write on paper. Where laptops are used the information seem unstructured, with clinicians having trouble finding the information they or my relative needs. Much of the care feels disjointed … Not once have records been shared with my relative, for checking detail or accuracy.”* **Carer 1727**

As well as concerns about documentation, respondents also questioned whether other health professionals could access their record at the point of need. Some raised concerns about their information being accessed by parties to which they had not granted permission.

> *“I have had my healthcare record shared with an insurance company by my gp without my permission … I have family members who do not know of my diagnosis and I do not wish [them] to know, so am worried that if this was a public message on my medical records it would be released without my permission.”* **LTC 194**

They were also aware that the existence of a digitally documented record did not mean health professionals would access it, and if they did that, they would provide care in line with documented wishes and preferences.

> *“My wife and I have registered our preferences / requirements with our GP and our local hospital, but we are both concerned that the information will be overlooked when the time arrives because hospital staff and particularly GP staff appear to have very poor understanding / knowledge of such advanced decisions.”* **TI 1074**

#### Theme 4: Preserving Dignity and Respect: Understanding individual contexts

Respondents described the type of information they considered important to document to inform their future care. Preferences that preserve dignity and respect were favoured over life-preserving medical interventions. It was particularly important to them that physical, psychological, social and spiritual needs are understood by family as well as health professionals. The type of preferences reported included the preferred place of care, remaining with assistance animals, dietary needs and how their specific health conditions should be managed. They considered the importance of stating treatments not yet available (such as a medically assisted death) in the case of these becoming a possibility.

> *“I have a spinal cord injury which requires very specialist care to manage bladder, bowels, pressure sore preventions, spasms, muscle contractions if legs are not stretched. I am extremely anxious as I currently manage my own care as GPs and local hospitals have virtually no knowledge … I will definitely want appropriate care to end my life with dignity.”* **LTC 1202**

Patients were aware of potential discrepancies between family members’ perceptions of how they should be cared for. They expressed concerns that future treatments and care may not align with stated wishes and preferences in families with different views. They saw a role for the digital record in guiding appropriate care by providing evidence of the patient’s preferences to family members.

> *“I had a major surgery so filled in an electronic record and printed it off … We have done the same for my elderly mother … the fact we have them makes me confident that things will go both how my mother wishes (my siblings were given copies of hers so there can be no arguments) and my wishes will be carried out.”* **LTC 593**

Participants talked about self-management of their health condition. Comments reflected their anxiety that information necessary to be able to do this in a way that preserves dignity, respect and comfort, would not be available to health professionals looking after them. Digital systems were perceived as a means of communicating information about preferred and holistic methods of symptom management, should the patient not be able to communicate this themselves.

> *“I worry about the practical care in hospital because of the daily pain I have; in the event I am not able to communicate to my caregivers I fear being manhandled.”* **LTC 1509**

They welcomed a documented plan that is accessible to health professionals, and they envisaged it would support having their wishes respected, either through supporting carers in advocating for patients or to be a substitute for advocacy for those with no next of kin.

> *“I personally am worried that my next of kin is not strong enough to advocate for me at end of life. I am therefore determined to document my wishes clearly to reduce risk of wishes not being followed through lack of courage or confidence.”* **Carer 941**

## Discussion

This large-scale survey of patients and the public captured experiences and views of how shared patient records support patients receiving palliative and end-of-life care. Respondents were in favour of their health and end-of-life wishes and preferences being shared across all settings but only had moderate levels of confidence that their information would be accessed or acted upon. Valued elements of digital advance care planning systems include detailing the people that a patient would want to involve in care decisions and preferences, alongside an ability to view, edit, and share their digital records with family. Perceptions of how digital systems facilitate the delivery of care that aligns with patient preferences are influenced by personal or observed experiences of how health and care were previously supported by the availability of patient information. Digital approaches are considered to benefit patients where participants already feel supported by these systems or where participants view them as a solution to avoiding previously encountered issues.

Low confidence that health professionals will read patient wishes and preferences aligns with research that highlights few hospital-based health professionals access information recorded by community-based health professionals and rarely use it for decision-making (28). Concerns about the uncertainty about where personal information was held, which services could view it, and a lack of confidence in its accuracy align with previous and recent research in the UK(21). Interviews with patients undertaken by Caine and colleagues (29) revealed that no participants expressed knowledge about how widely their health information was shared and half did not know what data was stored. In this study, participants valued the potential to view and edit their digital records online or via a mobile application. They were also keen that information could be shared with family members, subject to patient consent. This may provide crucial information to support caregiver decision-making in multiple ways. For example, there can be variable accuracy in predicting a partner’s end-of-life treatment preferences(30) and also scepticism from surrogate decision-makers about patient preferences (31). Furthermore, sharing information may circumvent both family pressures to continue treatment and the effect of grief on carers’ capacity to advocate can disrupt the care provision that is aligned with preferences(32). Our findings suggest that by accessing up-to-date patient information via a shared digital record, carers may feel supported when asked to make end-of-life decisions on behalf of the patient.

This study highlights the need to consider patient and caregiver perspectives on the content and implementation of digital systems for sharing health and care preferences at the end of life. Participants’ preferences for content were often informed by a need to preserve dignity and respect rather than extend life. Previous research resonates with the findings of this study relating to the importance of treatment preferences based on self-dignity and the value of living (32), and focussing on goals and values (33). Patient participants stated the importance of health professionals and family members’ understanding and acceptance of the rationale behind their choices, however, many were concerned that choices would be questioned and rejected if not understood. Freely available digital resources where patients enter information about personal values and preferences may provide assurance in the event of disagreement, particularly if a printed copy is retained (e.g. (15, 34)). Implemented well, online advance care planning tools can be practical and feasible (16, 35-38). The implementation of systems that support individual choice in the disclosure of their information should include easy patient access to their digital record, an overview of current sharing permissions, control over access, contextual privacy controls and notification when information is accessed[20].

## Strengths and limitations

To our knowledge, this is the first large-scale national survey to provide data about patient and carer views on how they perceive documentation and sharing personal health information, and wishes and preferences to support care at the end of life. As this was an online survey, we could not calculate response rates; however, we obtained a large sample size. Although there was a relatively low response from people with a terminal condition compared with other groups, the response from carers was high, and their comments indicated a breadth of experience caring for someone at the end of life. To preserve the anonymity and confidentiality of respondents and to keep the survey brief, neither demographic nor health information was gathered. This enabled us to maximise responses and the opportunities to capture a wide range of experiences, but we were not able to report patient or caregiver characteristics. In common with other large-scale cross-sectional healthcare experience surveys, negative experiences were more detailed and specific, providing richer material than positive experiences (39, 40). As a result, we may have missed out on detailed information about positive experiences and views of how digital systems are meeting patient needs. This could be why many comments focused on issues and concerns about discussing, documenting, and using advance care planning information, and the need for digital systems to provide better content and access to address these issues. Less information was provided about how digital systems had helped align care with patients’ wishes.

## Conclusion

Patients’ and carers’ views on digital documentation and sharing of information to support care at the end of life were shaped by prior experiences of how the availability of this information had supported care - or could have if it were available. Digital systems are increasingly being used to document health and advance care planning information. While patients and carers report the potential for digital systems to facilitate documentation and sharing of their information, they also question how these systems will support care in reality, and how accessible will they be to patients once created. Efforts should be made to build confidence and clarify the expectations of patients and members of the public around the documentation of their wishes and preferences for care alongside the subsequent sharing and use of this information. Furthermore, future research is required to explore whether such patient and carer access to their record influences confidence in the accuracy of content and the likelihood of care being delivered in line with their wishes.

## Supporting information

Supplemental Online Survey

## Data Availability

All data produced in the present study are available upon reasonable request to the authors

## Acknowledgements

We would like to acknowledge Compassion in Dying and the Professional Records Standards body for agreement to share their dataset.

## Authorship

JB conceived the study with support from MA. JB led the study design with support from all authors, including in the analysis and interpretation of data. JB drafted the initial manuscript, with revisions and critical input from all authors. All authors approved the final version for publication and have participated sufficiently in the work to take public responsibility for its content.

## Ethics approval

Ethics approval was sought from the University of Leeds School of Medicine Research Ethics Committee (reference: MREC 21-032). This study was conducted in accordance with the relevant guidelines and regulations. Participants indicated their agreement to participate via the online survey.

