## Supplemental Online Survey for "Patient and public experience and views on digital systems for sharing records for health and care preferences at the end of life"

**APPENDIX 1: Online Survey designed using Survey Monkey ©**

**Webpage 1 (introduction)**

This survey has seven questions and will take about five minutes to complete.

Any personal information you give us won’t be shared with anyone outside of the Professional Records Standards Body or Compassion in Dying.

We might share some of the things you say anonymously outside of our organisations to improve policy or practice.

**Webpage 2: PLEASE RATE HOW CONFIDENT YOU FEEL ABOUT THE FOLLOWING STATEMENTS.**

*Rating scale is 1 to 5, where 1 is not confident at all and 5 is very confident OR select “Don’t know”*

a. The healthcare team supporting or treating me will have access to the information about my health that they need when caring for me at the end of life.
b. The healthcare team supporting or treating me will have access to my end-of-life wishes and preferences.
c. The healthcare team supporting or treating me will have access to information on who I want to be involved in decisions about my health and care at the end of life.

**Webpage 3: WHAT WOULD GIVE YOU THE MOST CONFIDENCE THAT YOU OR A LOVED ONE WOULD GET THE CARE THAT’S RIGHT FOR YOU/ THEM AT THE END OF LIFE?**

***Please rate how much each statement matters to you.***

*Rating scale is 1 to 5, where 1 is this does not matter to me and 5 is this is very important to me OR select Don’t know*

a. The healthcare team supporting or treating me can see details of the people I want to be involved in decisions about my care.
b. The healthcare team supporting or treating me can see my preferences such as where I want to be cared for.
c. The healthcare team supporting or treating me can see which treatments I do and do not want.
d. I can view my end-of-life care record via a website or app.
e. I can record and make changes to my end-of-life care preferences via a website or app.
f. I can share my end-of-life care record with my family and loved ones.

**Optional comments box 1:** If there’s something else in relation to your end-of-life care record that is important to you, please tell us here.

**Webpage 4: WE WOULD LIKE TO KNOW HOW YOU FEEL ABOUT INFORMATION ABOUT YOUR HEALTH BEING SHARED WITH YOUR HEALTHCARE TEAM (FOR EXAMPLE YOUR DIAGNOSIS OR MEDICATIONS).**

***Please select which of the statements applies to you:***

1. I would like this information to be available to any healthcare professional supporting me whenever they need it, without me having to give permission for the information to be shared with them.
2. I would like to be asked once for my permission to share this information with any healthcare professional supporting me who needs it.
3. Neither

**Optional comments box 2** *Please tell us more*.

**Webpage 5: WE WOULD LIKE TO KNOW HOW YOU FEEL ABOUT YOUR END-OF-LIFE WISHES AND PREFERENCES BEING SHARED WITH YOUR HEALTHCARE TEAM (FOR EXAMPLE TREATMENTS YOU DO OR DO NOT WANT, THE PEOPLE YOU WOULD LIKE INVOLVED IN YOUR CARE, OR WHERE YOU WOULD LIKE TO BE CARED FOR).**

***Please select which of the statements applies to you.***

1. I would like this information to be available to any healthcare professional supporting me whenever they need it, without me having to give permission for the information to be shared with them.
2. I would like to be asked once for my permission to share this information with any healthcare professional supporting me who needs it.
3. Neither

**Optional comments box 3** *Please tell us more.*

**Webpage 6: WE WOULD LIKE TO BETTER UNDERSTAND HOW SHARING END-OF-LIFE RECORDS CAN IMPACT ON PEOPLE’S LIVES.**

***If you’ve had a specific experience, or have thoughts about your wishes being known at the end of life, could you tell us a bit more about it? We’d particularly like to hear from you if you:***

- *Have had a bad or worrying experience where your (or a loved one’s) end-of-life wishes were not available to the people who needed it.*
- *Have had a good experience where your information was shared with others in a way which meant you felt confident or received good care.*
- *Are worried or anxious about your wishes or healthcare information being known when it matters as you’re nearing the end of life.*
- **Optional Comments box 4**

**Webpage 7: WE WOULD LIKE TO UNDERSTAND HOW PEOPLE’S EXPERIENCES OF END-OF-LIFE CARE RECORDS VARIES ACROSS THE COUNTRY. IN WHICH AREA OF THE UNITED KINGDOM DO YOU LIVE?**

- **Please select one region (from list of UK counties)**
